# Population-based sequencing of *Mycobacterium tuberculosis* reveals how current population dynamics are shaped by past epidemics

**DOI:** 10.1101/2022.01.24.22269736

**Authors:** Irving Cancino-Muñoz, Mariana G. López, Manuela Torres-Puente, Luis M. Villamayor, Rafael Borrás, María Borrás-Máñez, Montserrat Bosque, Juan J. Camarena, Caroline Colijn, Ester Colomer-Roig, Javier Colomina, Isabel Escribano, Oscar Esparcia-Rodríguez, Francisco García-García, Ana Gil-Brusola, Concepción Gimeno, Adelina Gimeno-Gascón, Bárbara Gomila-Sard, Daminana González-Granda, Nieves Gonzalo-Jiménez, María Remedio Guna-Serrano, José Luis López-Hontangas, Coral Martín-González, Rosario Moreno-Muñoz, David Navarro, María Navarro, Nieves Orta, Elvira Pérez, Josep Prat, Juan Carlos Rodríguez, Ma. Montserrat Ruiz-García, Hermelinda Vanaclocha, Valencia Region Tuberculosis Working Group, Iñaki Comas

**Affiliations:** Tuberculosis Genomics Unit, Instituto de Biomedicina de Valencia (IBV-CSIC), 46010 Valencia, Spain; Unidad Mixta “Infección y Salud Pública” (FISABIO-CSISP), 46020 Valencia, Spain; Microbiology Service, Hospital Clínico Universitario, 46010 Valencia, Spain; Microbiology and Parasitology Service, Hospital Universitario de La Ribera, 46600 Alzira, Spain; Microbiology Service, Hospital Arnau de Vilanova, 46015 Valencia, Spain; Microbiology Service, Hospital Universitario Dr. Peset, 46017 Valencia, Spain; Department of Mathematics, Faculty of Science, Simon Fraser University, V5A 1S6 BC, Canada; Microbiology Laboratory, Hospital Virgen de los Lirios, 03804 Alcoy, Spain; Microbiology Service, Hospital de Denia, 03700 Denia, Spain; Bioinformatics and Biostatistics Unit, Centro de Investigaciones Príncipe Felipe, 46012 Valencia, Spain; Microbiology Service, Hospital Universitari i Politècnic La Fe, 46026 Valencia, Spain; Microbiology Service, Hospital General Universitario de Valencia, 46014 Valencia, Spain; Microbiology Service, Hospital General Universitario de Alicante, 03010 Alicante, Spain; Microbiology Service, Hospital General Universitario de Castellón, 12004 Castellón, Spain; Microbiology Service, Hospital Lluís Alcanyis, 46800 Xativa, Spain; Microbiology Service, Hospital General Universitario de Elche, 03203 Elche, Spain; Microbiology Service, Hospital Universitario de San Juan de Alicante, 03550 Alicante, Spain; Microbiology Service, Hospital de la Vega Baixa, 03314 Orihuela, Spain; Microbiology Service, Hospital San Francesc de Borja, 46702 Gandía, Spain; Subdirección General de Epidemiología y Vigilancia de la Salud y Sanidad Ambiental de Valencia (DGSP), 46020 Valencia, Spain; Microbiology Service, Hospital de Sagunto, 46520 Sagunto, Spain; CIBER of Epidemiology and Public Health (CIBERESP), Madrid, Spain

**Author notes:** corresponding authors: Mariana Gabriela López. Instituto de Biomedicina de Valencia, Calle Jaume Roig 11, 46010, Valencia, Spain., Iñaki Comas. Instituto de Biomedicina de Valencia, Calle Jaume Roig 11, 46010, Valencia, Spain. I. C-M and M.G.L contributed equally to this work.

**Keywords:** Tuberculosis, transmission, genomic epidemiology, whole-genome sequencing

## Abstract

**Background:** Transmission has been proposed as a driver of tuberculosis (TB) epidemics in high-burden regions, with negligible impact in low-burden areas. Genomic epidemiology can greatly help to quantify transmission in different settings but the lack of whole genome sequencing population-based studies has hampered its use to compare transmission dynamics and contribution across settings.

**Methods:** We generated an additional population-based sequencing dataset from Valencia Region, a low burden setting, and compared it with available datasets from different TB settings to reveal heterogeneity of transmission dynamics and its public health implications. We sequenced the whole genome of 785 *M. tuberculosis* strains and linked genomes to patient epidemiological data. We applied a pairwise distance clustering approach and phylodynamics methods to characterize transmission events over the last 150 years, in Valencia, Spain (low burden), Oxfordshire, United Kingdom (low burden) and a high-burden (Karonga, Malawi).

**Results:** Our results revealed high local transmission in the Valencia Region (47.4% clustering), in contrast to Oxfordshire (27% clustering), and similar to a high-burden setting like Malawi (49.8% clustering). By modelling times of the transmission events, we observed that settings with high transmission are associated with uninterrupted transmission of strains over decades, irrespective of burden.

**Conclusions:** Our results underscore significant differences in transmission between TB settings even with similar burdens, reveal the role of past epidemic in on-going TB epidemic and highlight the need for in-depth characterization of transmission dynamics and specifically-tailored TB control strategies.

**Funding:** European Research Council under the European Union’s Horizon 2020 research and innovation program (Grants 638553-TB-ACCELERATE, 101001038-TB-RECONNECT), and Ministerio de Ciencia e Innovación (Spanish Government, SAF2016-77346-R and PID2019-104477RB-I00)

## Introduction

Tuberculosis (TB) is one of the top 10 most deadly infectious diseases according to the World Health Organization (WHO). In 2019 were reported 10 million new TB cases and 1.4 million deaths, with these numbers likely to increase due to the COVID-19 pandemic (Glaziou, 2020). Recognizing heterogeneity across settings in the population-level dynamics of tuberculosis is key to advance to new stages in local and global TB control (Mathema et al., 2017). Recent transmission significantly contributes to the global TB-burden mostly in the high incidence regions and its control is imperative to achieve the goal of the End TB Strategy (Guerra-Assunção et al., 2015; “The transmission of Mycobacterium tuberculosis in high burden settings,” 2016). On the contrary, in many countries close to the pre-elimination phase (<5/100,000 cases) ongoing transmission plays a minor role and control strategies focused on latent TB infection (LTBI) mostly from imported cases (Menzies et al., 2018). However, whether burden can be used as a proxy of recent transmission is not clear and understanding transmission dynamics for each country is key for tailor-made strategies.

Measuring transmission is still challenging, it can be achieved by comparing the pathogen genomes of culture positive cases with some limitations, for example all transmission cases associated with LTBI or those with negative culture cannot be analyzed. However, it allows us to compare transmission clustering rates across countries in a standard way. Whole-genome sequencing (WGS) represents a widely applied tool in the study of TB epidemiology and transmission based on the pairwise single nucleotide polymorphisms (SNPs) distance (Gardy et al., 2011; “Whole-genome sequencing to delineate Mycobacterium tuberculosis outbreaks: a retrospective observational study,” 2013). WGS displays higher resolution, provides accurate results tracking recent transmission (“Aiming for zero tuberculosis transmission in low-burden countries,” 2017, “Role and value of whole genome sequencing in studying tuberculosis transmission,” 2019; Jajou et al., 2018; Meehan et al., 2019) and reports greater agreement with epidemiological results (Nikolayevskyy et al., 2016; Roetzer et al., 2013). Despite WGS reliability, there exists controversy regarding the SNP threshold employed to delineate genomic clusters. A cut-off of 5 SNPs has been widely accepted for the clustering of recently linked cases (Meehan et al., 2019; “Role and value of whole genome sequencing in studying tuberculosis transmission,” 2019) while an upper value of 12 SNPs also incorporates older transmission events (“Whole-genome sequencing to delineate Mycobacterium tuberculosis outbreaks: a retrospective observational study,” 2013); however, the extent to which the identification of those cases can aid epidemiological investigations remains controversial (Bjorn-Mortensen et al., 2016; Jajou et al., 2018). It is also unclear the extent to which those cutoffs apply to all settings given differences in social, host and pathogen factors across settings. Even if universal, understanding transmission dynamics goes beyond recent transmission events, which have an actionable value for public health, but that do not capture the long-term dynamics in a population.

The lack of WGS studies at the population level represents the main limitation to the validation of these thresholds across clinical settings and to understand the transmission dynamics in different settings. Here we use available datasets from a low burden setting (Oxfordshire, incidence 8.4 cases per 100,000) and from a high burden setting (Malawi, incidence 87 cases per 100,000) and compare to a newly generated dataset.

Spain is a low-incidence country (9.3/100,000) where the contribution of recent transmission to local TB burden remains largely unknown. We applied WGS to investigate the epidemiology and dynamics of TB transmission in the Valencia Region, the fourth most populated region of the country, over three years, and evaluated the general use of an SNP threshold in cluster definition in this particular setting. Compared with similar population-based studies from locations with different TB burdens (Guerra-Assunção et al., 2015; Walker et al., 2014), transmission in Valencia has a prominent role in the current epidemics, with a genomic clustering rate higher than the other low-burden and closer to high-burden settings. Furthermore, our results demonstrate that current TB incidence in Valencia and Malawi mainly derives from sustained transmission over time, with the majority of the linked cases currently observed coming from long-term transmission chains established around 30 years ago.

## Results

### *M. tuberculosis* population structure and demographic characteristics in Valencia Region

We sequenced 77% of the TB culture-positive cases reported between 2014-2016 in Valencia Region (Supplemental Table 1). 10 samples were removed as non-MTBC isolates or likely mixed infections (Supplemental figure 1). We identified 6 different lineages(L) circulating in the region (Coll et al., 2014; Stucki et al., 2016), with L4 the most frequent (92.1%) (Figure 1A).

**Table 1.** Dating of local genomic clusters (gCluster). Times of the oldest and youngest local gClusters obtained by a Bayesian analysis are presented, with values in years (AD) and 95% highest posterior density given in brackets. The number of gClusters and clustering percentage is provided for each dataset. The median distance ranges for all gClusters are also detailed.

| Dataset | Sampling period | Local samples | N local gCluster | Local clustering | Median distance range | Oldest gCluster | Youngest gCluster |
| --- | --- | --- | --- | --- | --- | --- | --- |
| Oxfordshire | 2006-2012 | 74 | 6 | 27% | 0-7 | 1993 [1982-2003] | 2009 [2003-2012] |
| Malawi | 2008-2010 | 106 | 40 | 49.80% | 0-14 | 1979 [1968-1988] | 2009 [2004-2010] |
| Valencia Region | 2014-2016 | 456 | 65 | 47.40% | 0-11 | 1985 [1972-1996] | 2015 [2012-2016] |

**Figure 1.**
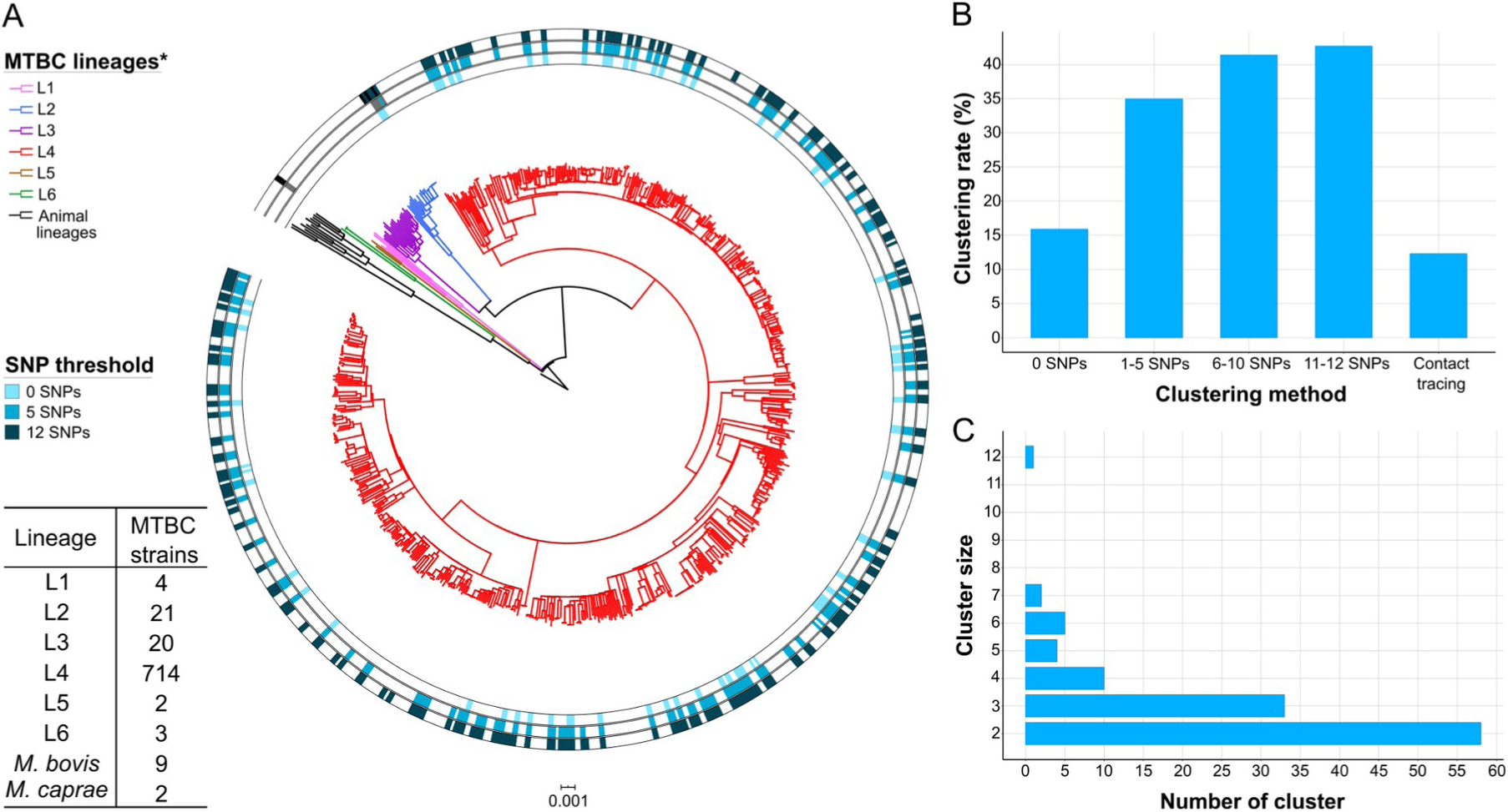
Genomic characterization of the study region. A. Phylogeny of 775 TB isolates collected during 2014-2016. Each ring represents genomic clusters detected by different SNP thresholds (0, 5, 10, and 12 SNPs). *M*.*canneti* was used as an outgroup. **B**. Clustering percentage, i.e. percentage of samples within clusters for different SNP thresholds. **C**. Number of genomic clusters by different cluster sizes. A 12-SNP threshold was used as a standard. Cluster sizes of 8 to 11 samples were not detected. *Nomenclature proposed by Comas et al. (Comas et al., 2013).

Characteristics of TB cases are summarized in Supplemental Table 2, reporting the sequenced samples as a representative subset of the total culture-positive cases. Detailed epidemiological analysis is presented in Supplemental Table 3, remarkably 63% of all cases were Spanish-born patients, while 30% came from high-incidence countries and 7% from other low-incidence countries. 14% of residents are foreign-born, thereby accounting for a TB incidence of 23.6 vs. 6.9 cases per 100,000 among Spanish-born patients. When observed risk factors, we found that 12.4% of patients suffered social exclusion, which was more prevalent among foreign-born patients (OR 3.1, CI 1.9-5.1, p<0.001). Diabetes was present in 10.4% of cases; although this was more prevalent in Spanish-born patients (OR 2.7, CI 1.5-5.4, p<0.001), values were similar to disease prevalence in the general population.

### Epidemiological and genomic clustering

Classic contact tracing identified 66 epidemiological clusters, including 97 cases, accounting for 12.5% of transmission in the Valencia Region (Figure 1B). Spanish-born and foreign-born patients equally formed part of an epidemiological cluster. Considering a pairwise distance threshold of 12 SNPs, we identified 112 genomic clusters, including 331 (42.7%) patients, with clusters including from 2 to 12 cases (Figure 1C, Supplemental Table 4). Although these clusters included foreign-born patients, Spanish-born patients were more likely part of genomically-linked groups (OR 2, CI 1.44-2.79, p<0.001). In this regard, 42 genomic clusters exclusively comprised Spanish-born patients and 8 included only foreign-born patients. Besides Spanish origin and pulmonary localization of TB (OR 2.5, CI 1.60-3.98, p<0.005), no social or risk factor appeared associated with transmission (Supplemental Table 3). In addition, 90% of TB cases in Valencia Region are susceptible to all antibiotics used in treatment, so resistance mutations do not have an impact in the clustering.

We also assessed genomic clusters considering different SNP thresholds, and observed that independently of the cut-off considered, the clustering rate obtained by contact tracing was always lower than the genomic estimates (Figure 1B). A high number of genomic links were not detected by epidemiological inspection, while some epidemiological links were not corroborated by any genomic clustering threshold (Figure 2A). Comparison of both approaches revealed that only 15.4% of the 331 patients within genomic clusters (12 SNPs) had an identified epidemiological link (Supplemental Results, Supplemental Table 5).

**Figure 2.**
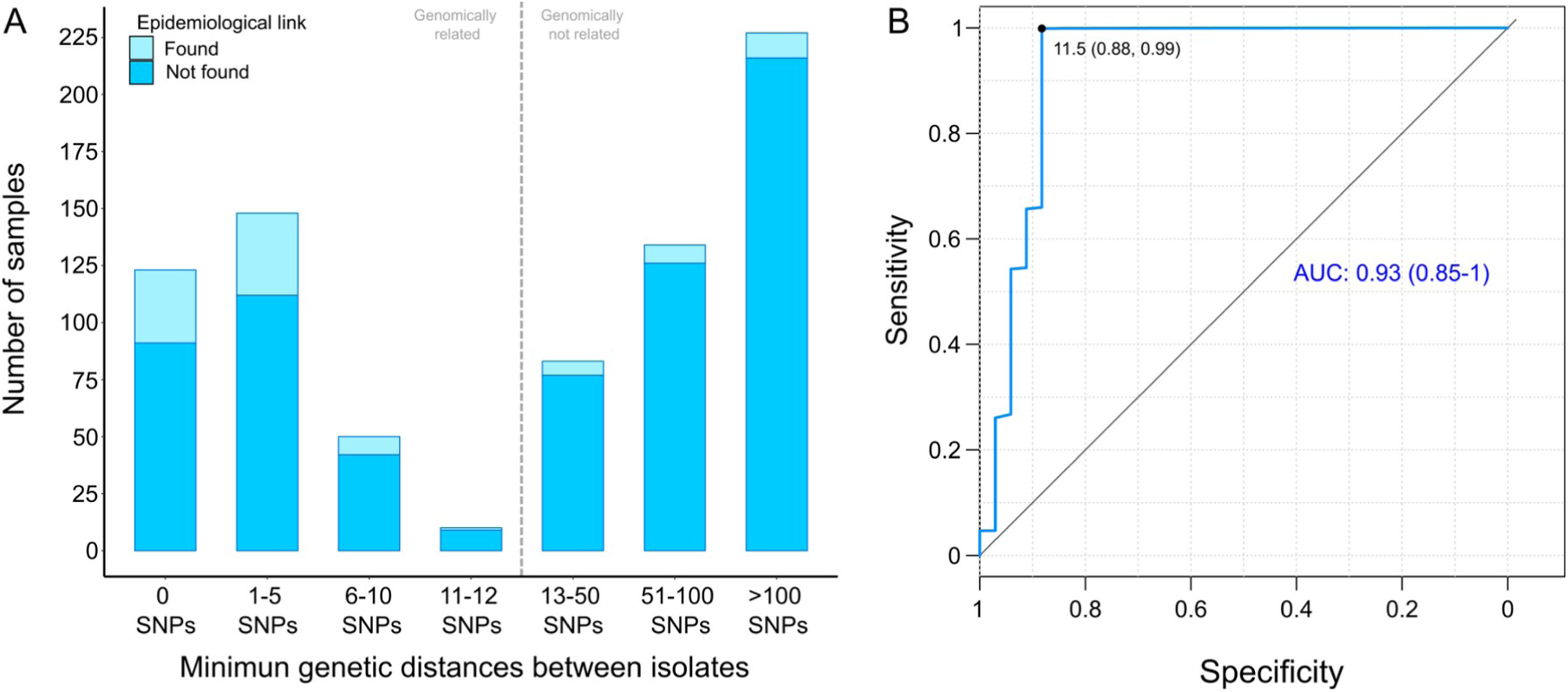
Comparison between epidemiological and genomic clustering. **A**. Clustered samples using different pairwise distance thresholds, bars denote the number of cases within clusters for each SNP threshold. Grey dashed line separates the genomically linked samples (clustered) from those unlinked. **B**. ROC curve for different pairwise distance thresholds between 0 and 2,000 SNPs, indicating the optimal SNP cut-off values with its correspondent specificity and sensitivity values, the area under the curve (AUC), and its confidence intervals.

We benchmarked WGS as a tool to quantify transmission against contact tracing (Diel et al., 2019), using the latter as the gold standard (Supplemental Table 6). In general, as the SNP threshold decreases, sensitivity diminishes, but specificity and accuracy increase. By a ROC curve, we established 11.5 SNPs as the optimal value for the SNP cut-off that maximizes the agreement between epidemiological investigation and genomic data, and genomic clustering appears as an adequate approach to discriminate transmission, as the area under the curve is higher than 0.9 (Figure 2B). Then, we used 12 SNPs threshold to define clusters in the following analyses.

### Genetic thresholds for transmission are not universal across settings

We calculated the percentage of Spanish-born cases clustered by a range of pairwise distances (0-150 SNPs) and compared with the clustering of local cases in other settings (Guerra-Assunção et al., 2015; Walker et al., 2014), where most of the 70% of all culture-positive cases were sequenced. We observed a bimodal pattern for Oxfordshire, with the transmission groups clearly differentiated from the other unlinked cases with distances higher than 50 SNPs. These findings agree with the 12-SNP value proposed as a means to identify transmission in datasets from low-burden countries (Walker et al., 2014). For the Valencia Region and Malawi, strains group in a large range of distance thresholds (SNPs 0-150). Thus, there exists a continuous clustering throughout the distance values. The results strongly suggest that a strict transmission threshold of 12 SNPs (or any other threshold) does not apply to all settings, particularly those with higher transmission burdens (Figure 3A) and particularly if we want to understand long-term transmission dynamics.

**Figure 3.**
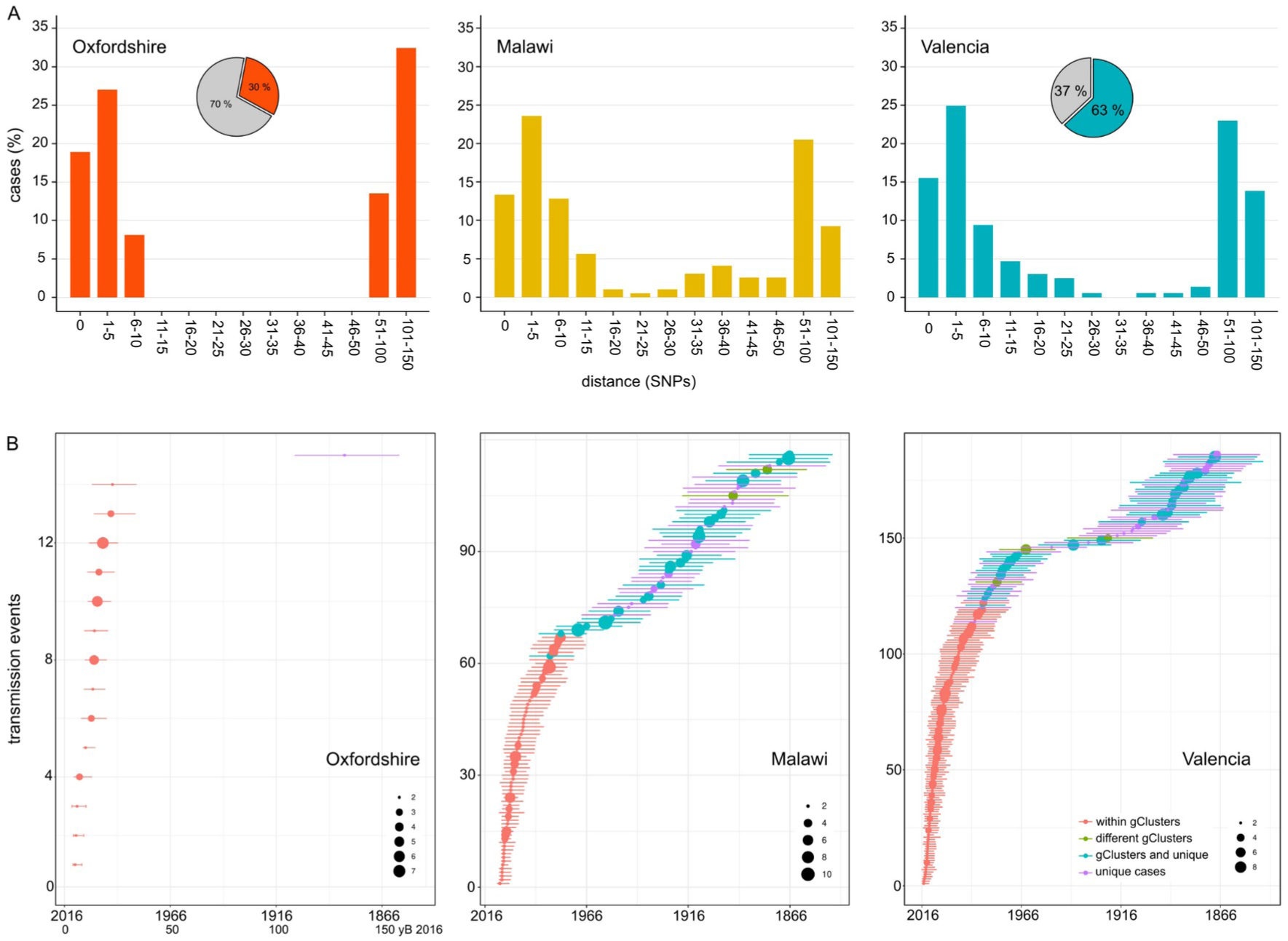
Transmission dynamics analysis. **A**. Distribution of Spanish-born cases clustered by different pairwise distance SNP thresholds. Cases are expressed as the percentage of the plotted samples. Pie charts represent the proportion of Spanish-born (color) and foreign-born (gray) cases in each dataset. **B**. Age of local transmission events over time in each setting. Circles represent median time, and lines 95% high probability density for each transmission event counted. Circle size represents the number of samples included in the corresponding event. Red denotes those transmission events including only samples within the same genomic transmission clusters (gClusters), green denotes events involving samples from different gClusters, blue denotes samples within gClusters and unique, and purple denotes unique cases.

### Age of local genomic clusters at different SNP thresholds and impact on public health

Next, we evaluated how old are the genomic clusters identified by the standard 12 SNP threshold. Thus, we inferred the age of the local genomic clusters (gClusters) for the three settings. Dating results of the youngest and the oldest gClusters are summarized in Table 1, while complete results are detailed in Supplemental Tables 7-9. We can trace gClusters 31 years back from the most recent sample collected for both the Valencia Region and Malawi; however, we only retrieved samples that formed part of gClusters, 19 years before the most recent Oxfordshire sample. The alternative calibration samples included (Supplemental Methods) displayed similar results, thereby allowing comparisons among datasets. Thus many gClusters based on 12 SNP thresholds are beyond the action of public health interventions. In fact, when looking at epidemiological linked cases in the Valencia Region, most of them have a common ancestor less than 10 years before the most recent sample, and the distance between samples typically ranged between 0-4 SNPs, with only one cluster separated by 11 SNPs (Supplemental Table 5). While the ROC curve indicated a 12 SNP threshold to capture most epidemiological links the reality is that strains linked by more than 5 SNP are beyond the action of public health interventions as they involve too old transmission links. Our results imply that events useful for public health investigations are better captured by a 5 SNP threshold even though some epidemiological links are missing. But the reverse is also true, and more dramatic. Even when using a 5 SNP threshold public health only identifies around 15% of the cases in genomic clusters. This holds true even for pairs of isolates with 0 SNP differences. As seen in high-burden countries, when transmission has a prominent role, many transmission events occur outside the traditional household or work settings.

### Transmission events over time and between clinical settings highlight distinct epidemic dynamics

In order to evaluate transmission dynamics over time, we traced transmission events back to 150 years before 2016 (yB 2016) by using genomic data from local-born patients to avoid the influence of imported genotypes. In the case of Oxfordshire, we identified 14 events between 5-25 yB 2016, with the next transmission event being inferred between 100-150 yB 2016 (Figure 3B, Supplemental figure 2, Supplemental Table 10). Thus, a gap of 75 years occurs between the most recent and the oldest transmission events, explaining why the 12 SNP threshold performs well in this setting as a transmission marker. However, even in this setting, genomic transmission clusters defined by a 12 SNP threshold can be traced back to up to 19 years (Table 1), which calls into question whether the 12 SNPs represent recent transmission in some cases. In the case of Malawi, we counted 70 events dating back 50 yB 2016 and 46 dated between 50-150 yB 2016 (Figure 3B, Supplemental figure 3, Supplemental Table 11). For the Valencia Region, we counted 143 events dated back 50 yB 2016 and 43 between 50-150 yB 2016 (Figure 3B, Supplemental figure 4, Supplemental Table 12). The gap detected in Oxfordshire is not observed in Malawi or Valencia.

The sampling of strains that shared a link decades ago in the Valencia Region can be explained in two ways; that the uninterrupted transmission of those strains until today or that the cases represent the progression of decades-old (latent) infections. We reasoned that if old reactivations contribute to strains in the Valencia region sampled during 2014-2016, we should see an increment in the age of the TB patients belonging to the older clusters (i.e., patients infected 20 years ago and have reactivated recently). We found no difference when comparing the age of the patients belonging to a gCluster with the inferred age of the cluster, (Welch two-samples t-test, p-values > 0.1, Supplemental figure 5, Supplemental Table 13), suggesting that the strains included in this study do not represent reactivations, and that uninterrupted transmission is the most likely explanation for the old links observed.

## Discussion

Here, we present the first national population-based study in the Valencia Region. We sequenced the whole genome of a representative proportion of all the TB notified cases that provides an accurate picture of the bacterial population structure, during three years. We exhaustively researched TB transmission linked to local epidemiological data and, by comparing to other settings, highlighted four main characteristics defining dynamics and influence on TB incidence.

### (I) Transmission can play a significant role in low*-*burden countries, *especially among* local-born patients

The percentage of genomically-linked cases (12 SNPs) of around 43% increases to 47% among the Spanish-born population -being 31% among imported cases-, suggesting that transmission among locally-born patients majorly contributes to burden. Percentages remain high when considering a stricter threshold of 5 SNPs for clustering (35% and 39%, respectively). We found higher transmission in the Valencia Region when compared to other low-burden settings, where clustering ranged between 14-16% (Jajou et al., 2018; Walker et al., 2014) and somewhat closer to that reported in high-burden TB countries (39-66%) (Guerra-Assunção et al., 2015; López et al., 2020). While high transmission burden in Valencia is associated with higher disease incidence in Spanish-born, reactivation of infections in imported cases from high-burden settings seem to be the significant drivers in other low-burden settings (Jajou et al., 2018; Kamper-Jørgensen et al., 2012; Walker et al., 2014). Thus our results highlight the heterogeneity of the TB epidemic even among countries with similar burden.

### (II) Community transmission majorly contributes to transmission burden

High genomic clustering suggests that many infections occurred outside the traditional household or work environment. In high-burden countries, which suffer from rampant community transmission (“The transmission of Mycobacterium tuberculosis in high burden settings,” 2016), epidemiological links are only identified in 18% of all genomically-clustered cases (Yang et al., 2018), a similar value to that observed in Valencia (15.4%), despite contact tracing occurring in 78% of cases. In those settings, studies have suggested that contact tracing among close contacts will not have a significant effect on TB incidence at a community level (McCreesh and White, 2018; Surie et al., n.d.), as transmission associates more with social drivers (Mathema et al., 2017). This likely explains the lack of agreement between genomic and epidemiologic clusters observed in the Valencia Region (62%) compared to other low-burden settings(Diel et al., 2019; Walker et al., 2014).

### (III) Genomic links are older than epidemiological links

The Valencia Region’s oldest genomic clusters dated to around 30 years before the sampling period. When considering only strains epidemiologically-linked, the oldest MRCA can be traced less than 10 years. Thus, a 12 SNP threshold identifies both recent and older transmission events. A 5 SNP threshold dates clusters between 1999-2015 in agreement with recent transmission rendering more actionable results for public health. However, a 5 SNP threshold still misses a percentage of cases linked by epidemiological data and vice versa, highlighting transmission complexity and the relevance of understanding its dynamics in each setting. Thus, a strict threshold has limitations and communicating a range, incorporating degrees of confidence, will be more valuable for public health interventions. This is particularly true in settings where transmission still has a prominent role.

### (IV) Continuous pairwise genetic distance distributions reflect decades-old transmission chains

The evaluation of local-born cases in the Valencia Region revealed continuous clustering across genetic distances, similar to Malawi. In both settings, differentiation between linked and unlinked cases seems arbitrary, as a clear SNP cut-off to delineate genomic transmission could not provide precise results (Figure 4A). This contrasts with the results of Oxfordshire, where clustering does not change in the range of 12-150 SNPs (Figure 4B). In this sense, the SNP threshold choice used to differentiate transmission from unrelated cases remains challenging even in low-burden settings and provides only tentative information (Meehan et al., 2019). An in-depth evaluation of clustering is needed to understand the particular transmission dynamics. Furthermore, the Valencia Region and Malawi also display continuous and sustained transmission events over time (Figure 4C). Those events outside the genomic transmission clusters likely reflect older contagion chains that still contribute to TB incidence today, as a consequence, clustering is continuous in settings exhibiting this transmission dynamics. The lack of effective past efforts to halt transmission may represent a plausible explanation. Epidemiological data demonstrates that Spain will likely attain a country profile similar to the UK and other low-burden, high-immigration countries. The higher transmission and the older age of transmission chains likely reflects a situation in which Spain suffered from higher disease incidence for most of the 20^th^ century, reflecting its lower socioeconomic status than neighboring countries. The current control strategies in place in the Valencia Region meet the WHO’s targets to reduce TB, including active case findings of close contacts since the 1990s. Improve TB control has led to a continuous drop in case numbers and to an incidence from 22 to 6.4 in the last 20 years. By contrast, Oxfordshire displays a bimodal distribution of clustering across pairwise distances, and also lacked transmission events other than those involving 12-SNP genomic clusters (Figure 4). These results agree with the robust reduction in both disease incidence and transmission that occurred until the beginning of the 1990s in the UK; after that, increased HIV infections, immigration and the emergence of TB drug resistance fueled the expansion of the non-eradicated TB (Glaziou et al., 2018). In accordance with this data, we dated ongoing transmission in Oxfordshire back to 1993.

**Figure 4.**
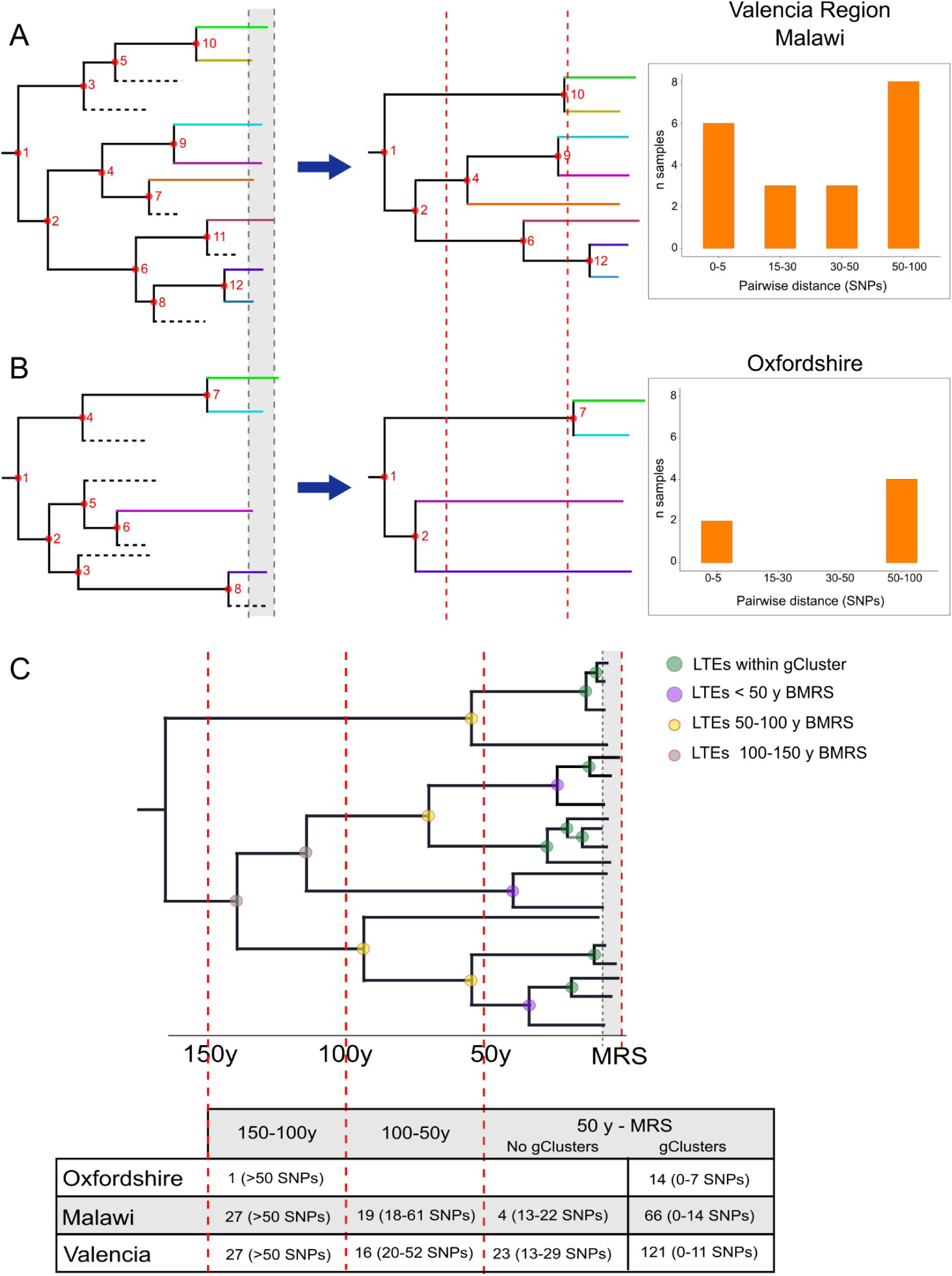
Hypothetical time trees indicating transmission events. **A**. (*Left)* The complete phylogeny, including all bacterial isolates and displaying multiple and sustained transmission events (nodes), over time. This scenario allows the reconstruction of a tree (*middle*) with several tips and multiple transmission events. A continuous distribution of clustered cases by different pairwise distances is retrieved (*right*) as observed in the Valencia Region and Malawi. **B**. A complete phylogeny (*left*) in which transmission is either too old or recent and few (or no) transmission events occurred in the middle time, led to the reconstruction of a tree (*middle*) in which few samples reach the present and fewer nodes are observed all over the tree. This scenario provides a bimodal distribution of clustered cases by pairwise distance (*right*) as observed for Oxfordshire. **C**. Time tree with local transmission events (LTEs) over time before the most recent sample (BMRS). The table (*bottom*) shows the number of events counted in each time period and the median distance range among the samples within the events for the three settings analyzed. For the period between the most recent sample (MRS) and 50yBMRS, events within (gClusters) and outside genomic clusters (No gClusters) are indicated. Vertical red lines indicate periods of time, horizontal dashed lines indicate missing samples, shaded areas indicate sampling period, and circles indicate transmission events with colors specified in the legend.

The main limitations of our analysis are inherent to the methodology, since only cases with positive cultures are sequenced. For the Valencia Region, cases included are an accurate representation of the epidemiological characteristics of the populations under study. Transmission is oversimplified by considering nodes as transmission events, while most transmissions will map to branches rather than nodes. However, knowing the exact timing of transmission is only possible for recent events and a proportion of cases (Xu et al., 2019), and not relevant for our comparative study which focuses mainly on old transmission events. Differences in the absolute number of cases in each dataset are irrelevant for comparison, since they all represent population-based studies with the same time-window sampling, thus the majority culture positive cases were included in the analysis. In this sense, the distribution of cases in clusters likely reflects the whole transmission dynamics of the settings.

Our results underscore a primary role for continuous transmission rather than LTBI reactivation or immigration in fueling TB incidence in the Valencia Region, as occurs in many high-burden settings (Bjorn-Mortensen et al., 2016; Guerra-Assunção et al., 2015; López et al., 2020; Yang et al., 2018). The opposite scenario occurs in other low-burden countries (Jajou et al., 2018; Walker et al., 2014) where transmission is limited and immigration from high-burden countries, also involving reactivation of the disease, represents the primary driver of incidence. In addition, reported meta-analysis from historical epidemiological studies suggests that, contrary to current assumptions, MTB infection may not be lifelong, and most people are able to clear it (Behr et al., 2019). This further suggests that the prevalence of LTBI is much lower than assumed, and most of the TB cases we see today are coming either from recent contagion or imported depending on the TB setting. Our data highlight how low-burden TB locations can entail very distinct scenarios that require specifically-tailored management, and that general TB guidelines should not be applied to all areas based solely on incidence rate (Lönnroth et al., 2015). Understanding heterogeneities in TB transmission dynamics is essential to define tailor-made interventions to halt transmission with a population-level impact, which is key to reducing the incidence of TB worldwide.

## Materials and Methods

Extended and detailed methods in Supplemental Information

### Sample selection and study design

1,388 TB cases were reported between 2014-2016 by the Valencian Regional Public Health Agency (DGSP), 1,019 with positive culture. All the available (785) samples were collected from 18 regional hospitals (Supplemental figure 1). Demographic, clinical, and microbiological records were obtained from the routine TB surveillance system, for 724 of the total samples. All diagnosed TB-positive patients completed a standardized questionnaire provided by the DGSP. *M. tuberculosis* structure and clustering analysis were performed with the total sequences. Epidemiological and transmission dynamics analysis were carried on with the samples with available information (724).

Approval for the study was given by the Ethics Committee for Clinical Research from the Valencia Regional Public Health Agency (*Comité Ético de Investigación Clínica de la Dirección General de Salud Pública y Centro Superior de Investigación en Salud Pública*). Informed consent was waived on the basis that TB detection forms part of the regional compulsory surveillance program of communicable diseases. All personal information was anonymized, and no data allowing patient identification was retained.

### DNA extraction and sequencing

Clinical isolates were cultured in Middlebrook 7H11 agar plates supplemented with 10% OADC (Becton-Dickinson) for three weeks at 37°C. After an inactivation step (90 °C, 15 min), DNA was extracted using the cetyl trimethyl ammonium bromide method from a representative sample from each patient (four-time plate scraping). All procedures were conducted in a Biological Safety Level 3 laboratory under WHO protocol recommendations. Sequencing libraries were constructed with a Nextera XT DNA library preparation kit (Illumina, San Diego, CA), following the manufacturer’s instructions. Sequencing was performed using the Illumina MiSeq platform.

### Bioinformatics Analysis

Data analysis was carried out following a validated previously-described pipeline (http://tgu.ibv.csic.es/?page_id=1794, (Meehan et al., 2019). Sequencing reads were trimmed with fastp (Chen et al., 2018), and kraken software (Wood and Salzberg, 2014) was then used to remove non-*Mycobacterium tuberculosis* complex (MTBC) reads. Filtered reads were mapped to an inferred MTBC common ancestor genome (https://doi.org/10.5281/zenodo.3497110) using BWA (Li and Durbin, 2009). SNPs were called with SAMtools (Li, 2011) and VarScan2 (Koboldt et al., 2012). GATK HaplotypeCaller (McKenna et al., 2010) was used for calling InDels. SNPs with a minimum of 10 reads (20X) in both strands and minimum base quality of 25 were selected and classified based on their frequency in the sample as fixed (>90%) or low frequency (10–89%). InDels with less than 20X were discarded. SnpEff was used for SNP annotation using the H37Rv annotation reference (AL123456.2). Finally, SNPs falling in genes annotated as PE/PPE/PGRS, ‘maturase,’ ‘phage,’ ‘13E12 repeat family protein’; those located in insertion sequences; those within InDels or in higher density regions (>3 SNPs in 10 bp) were removed due to the uncertainty of mapping. Next, variants were compared with recently published catalogues with validated association between mutations and phenotypic resistance (Ngo and Teo, 2019) in order to predict high-confidence resistance profiles to first- and second-line drugs. Lineages were determined by comparing called SNPs with specific phylogenetic positions established (Coll et al., 2014; Stucki et al., 2016). An in-house R script was used to detect mixed infections based on the frequency of lineage- and sublineage-specific positions (López et al., 2020). Read files were deposited in the European Nucleotide Archive (ENA) under the bioproject numbers PRJEB29604 and PRJEB38719 (Supplemental Table 1). Sequences from two population-based studies in Oxfordshire (Walker et al., 2014), with 92% of culture-positive cases sequenced, and Malawi (Guerra-Assunção et al., 2015), with 72% of culture-positive cases sequenced, were downloaded from ENA and analyzed as for the sequences generated in this study. All the custom scripts used are available in https://gitlab.com/tbgenomicsunit.

### Genomic clustering and phylogenetic analyses

The pairwise SNP distance was computed with the R *ape* package. Genomic clusters were constructed if the genetic distance between at least two patients’ isolates fell below a defined threshold. Cluster monophyly was confirmed in a maximum likelihood tree (50,184 SNPs). Timed phylogenies were inferred with Beast v2.5.1 (Bouckaert et al., 2014). Ancient TB DNA (Bos et al., 2014) and samples from a recent Spanish outbreak were included as calibration data. Dating was performed using GTR + GAMMA substitution model, a strict molecular clock model, and a coalescent constant size demographic model, as previously described (López et al., 2020). Three independent runs of Markov Chain Monte-Carlo length chains of 10 million were performed. Adequate mixing, convergence and sufficient sampling were assessed in Tracer v1.6, after a 10% burn-in.

### Tracking local transmission events over time

Transmission events were defined as nodes occurring over time phylogenies (Supplemental figure 6). The rationale for this approach is based on the assumption that if few pathogen mutations are expected to be observed during a host’s infection, as is the case of *M. tuberculosis*, lineages split only at transmission (Hall et al., 2016). To estimate the number of local transmission events, all ancestral nodes were counted, including local-born tips occurring within 150 years before 2016.

## Data Availability

Sequences were upload to ENA under the project numbers PRJEB29604 and PRJEB38719. Sequences detail, and associated information is detailed in Supplemental Table 1

## Notes

### Competing Interest Statement

The authors have declared no competing interest.

### Clinical Trial

This manuscript does not involve clinical trials

### Funding Statement

This project received funding from the European Research Council under the European Union's Horizon 2020 research and innovation program (Grants 638553-TB-ACCELERATE, 101001038-TB-RECONNECT) and by projects SAF2016-77346-R and PID2019-104477RB-I00 from Ministerio de Ciencia (Spanish Government).

### Author Declarations

Approval for the study was given by the Ethics Committee for Clinical Research from the Valencia Regional Public Health Agency (Comité Ético de Investigación Clínica de la Dirección General de Salud Pública y Centro Superior de Investigación en Salud Pública). Informed consent was waived on the basis that TB detection forms part of the regional compulsory surveillance program of communicable diseases.

## References

Aiming for zero tuberculosis transmission in low-burden countries. 2017.. The Lancet Respiratory Medicine 5:846–848.

Behr MA, Edelstein PH, Ramakrishnan L. 2019. Is infection life long? BMJ 367:l5770.

Bjorn-Mortensen K, Soborg B, Koch A, Ladefoged K, Merker M, Lillebaek T, Andersen AB, Niemann S, Kohl TA. 2016. Tracing Mycobacterium tuberculosis transmission by whole genome sequencing in a high incidence setting: a retrospective population-based study in East Greenland. Sci Rep 6:33180.

Bos KI, Harkins KM, Herbig A, Coscolla M, Weber N, Comas I, Forrest SA, Bryant JM, Harris SR, Schuenemann VJ, Campbell TJ, Majander K, Wilbur AK, Guichon RA, Wolfe Steadman DL, Cook DC, Niemann S, Behr MA, Zumarraga M, Bastida R, Huson D, Nieselt K, Young D, Parkhill J, Buikstra JE, Gagneux S, Stone AC, Krause J. 2014. Pre-Columbian mycobacterial genomes reveal seals as a source of New World human tuberculosis. Nature. doi:10.1038/nature13591

Bouckaert R, Heled J, Kühnert D, Vaughan T, Wu C-H, Xie D, Suchard MA, Rambaut A, Drummond AJ. 2014. BEAST 2: a software platform for Bayesian evolutionary analysis. PLoS Comput Biol 10:e1003537.

Chen S, Zhou Y, Chen Y, Gu J. 2018. fastp: an ultra-fast all-in-one FASTQ preprocessor. Bioinformatics 34:i884–i890.

Coll F, McNerney R, Guerra-Assunção JA, Glynn JR, Perdigão J, Viveiros M, Portugal I, Pain A, Martin N, Clark TG. 2014. A robust SNP barcode for typing Mycobacterium tuberculosis complex strains. Nat Commun 5:4812.

Comas I, Coscolla M, Luo T, Borrell S, Holt KE, Kato-Maeda M, Parkhill J, Malla B, Berg S, Thwaites G, Yeboah-Manu D, Bothamley G, Mei J, Wei L, Bentley S, Harris SR, Niemann S, Diel R, Aseffa A, Gao Q, Young D, Gagneux S. 2013. Out-of-Africa migration and Neolithic coexpansion of Mycobacterium tuberculosis with modern humans. Nat Genet 45:1176–1182.

Diel R, Kohl TA, Maurer FP, Merker M, Walter KM, Hannemann J, Nienhaus A, Supply P, Niemann S. 2019. Accuracy of whole-genome sequencing to determine recent tuberculosis transmission: an 11-year population-based study in Hamburg, Germany. Eur Respir J 54. doi:10.1183/13993003.01154-2019

Gardy JL, Johnston JC, Ho Sui SJ, Cook VJ, Shah L, Brodkin E, Rempel S, Moore R, Zhao Y, Holt R, Varhol R, Birol I, Lem M, Sharma MK, Elwood K, Jones SJM, Brinkman FSL, Brunham RC, Tang P. 2011. Whole-genome sequencing and social-network analysis of a tuberculosis outbreak. N Engl J Med 364:730–739.

Glaziou P. 2020. Predicted impact of the COVID-19 pandemic on global tuberculosis deaths in 2020. medRxiv 2020.04.28.20079582.

Glaziou P, Floyd K, Raviglione M. 2018. Trends in tuberculosis in the UK. Thorax.

Guerra-Assunção JA, Crampin AC, Houben R, Mzembe T, Mallard K, Coll F, Khan P, Banda L, Chiwaya A, Pereira RPA, McNerney R, Fine PEM, Parkhill J, Clark TG, Glynn JR. 2015. Large-scale whole genome sequencing of M. tuberculosis provides insights into transmission in a high prevalence area. eLife. doi:10.7554/elife.05166

Hall MD, Woolhouse MEJ, Rambaut A. 2016. Using genomics data to reconstruct transmission trees during disease outbreaks. Rev Sci Tech 35:287–296.

Jajou R, de Neeling A, van Hunen R, de Vries G, Schimmel H, Mulder A, Anthony R, van der Hoek W, van Soolingen D. 2018. Epidemiological links between tuberculosis cases identified twice as efficiently by whole genome sequencing than conventional molecular typing: A population-based study. PLoS One 13:e0195413.

Kamper-Jørgensen Z, Andersen AB, Kok-Jensen A, Bygbjerg IC, Andersen PH, Thomsen VO, Kamper-Jørgensen M, Lillebaek T. 2012. Clustered tuberculosis in a low-burden country: nationwide genotyping through 15 years. J Clin Microbiol 50:2660–2667.

Koboldt DC, Zhang Q, Larson DE, Shen D, McLellan MD, Lin L, Miller CA, Mardis ER, Ding L, Wilson RK. 2012. VarScan 2: somatic mutation and copy number alteration discovery in cancer by exome sequencing. Genome Res 22:568–576.

Li H. 2011. A statistical framework for SNP calling, mutation discovery, association mapping and population genetical parameter estimation from sequencing data. Bioinformatics 27:2987–2993.

Li H, Durbin R. 2009. Fast and accurate short read alignment with Burrows-Wheeler transform. Bioinformatics 25:1754–1760.

Lönnroth K, Migliori GB, Abubakar I, D’Ambrosio L, de Vries G, Diel R, Douglas P, Falzon D, Gaudreau M-A, Goletti D, González Ochoa ER, LoBue P, Matteelli A, Njoo H, Solovic I, Story A, Tayeb T, van der Werf MJ, Weil D, Zellweger J-P, Abdel Aziz M, Al Lawati MRM, Aliberti S, Arrazola de Oñate W, Barreira D, Bhatia V, Blasi F, Bloom A, Bruchfeld J, Castelli F, Centis R, Chemtob D, Cirillo DM, Colorado A, Dadu A, Dahle UR, De Paoli L, Dias HM, Duarte R, Fattorini L, Gaga M, Getahun H, Glaziou P, Goguadze L, Del Granado M, Haas W, Järvinen A, Kwon G-Y, Mosca D, Nahid P, Nishikiori N, Noguer I, O’Donnell J, Pace-Asciak A, Pompa MG, Popescu GG, Robalo Cordeiro C, Rønning K, Ruhwald M, Sculier J-P, Simunović A, Smith-Palmer A, Sotgiu G, Sulis G, Torres-Duque CA, Umeki K, Uplekar M, van Weezenbeek C, Vasankari T, Vitillo RJ, Voniatis C, Wanlin M, Raviglione MC. 2015. Towards tuberculosis elimination: an action framework for low-incidence countries. Eur Respir J 45:928–952.

López MG, Dogba JB, Torres-Puente M, Goig GA, Moreno-Molina M, Villamayor LM, Cadmus S, Comas I. 2020. Tuberculosis in Liberia: high multidrug-resistance burden, transmission and diversity modelled by multiple importation events. Microbial Genomics 6:e000325.

Mathema B, Andrews JR, Cohen T, Borgdorff MW, Behr M, Glynn JR, Rustomjee R, Silk BJ, Wood R. 2017. Drivers of Tuberculosis Transmission. J Infect Dis 216:S644– S653.

McCreesh N, White RG. 2018. An explanation for the low proportion of tuberculosis that results from transmission between household and known social contacts. Sci Rep 8:1–9.

McKenna A, Hanna M, Banks E, Sivachenko A, Cibulskis K, Kernytsky A, Garimella K, Altshuler D, Gabriel S, Daly M, DePristo MA. 2010. The Genome Analysis Toolkit: a MapReduce framework for analyzing next-generation DNA sequencing data. Genome Res 20:1297–1303.

Meehan CJ, Goig GA, Kohl TA, Verboven L, Dippenaar A, Ezewudo M, Farhat MR, Guthrie JL, Laukens K, Miotto P, Ofori-Anyinam B, Dreyer V, Supply P, Suresh A, Utpatel C, van Soolingen D, Zhou Y, Ashton PM, Brites D, Cabibbe AM, de Jong BC, de Vos M, Menardo F, Gagneux S, Gao Q, Heupink TH, Liu Q, Loiseau C, Rigouts L, Rodwell TC, Tagliani E, Walker TM, Warren RM, Zhao Y, Zignol M, Schito M, Gardy J, Cirillo DM, Niemann S, Comas I, Van Rie A. 2019. Whole genome sequencing of Mycobacterium tuberculosis : current standards and open issues. Nat Rev Microbiol 17:533–545.

Menzies NA, Cohen T, Hill AN, Yaesoubi R, Galer K, Wolf E, Marks SM, Salomon JA. 2018. Prospects for Tuberculosis Elimination in the United States: Results of a Transmission Dynamic Model. Am J Epidemiol 187:2011–2020.

Ngo T-M, Teo Y-Y. 2019. Genomic prediction of tuberculosis drug-resistance: benchmarking existing databases and prediction algorithms. BMC Bioinformatics 20:1–9.

Nikolayevskyy V, Kranzer K, Niemann S, Drobniewski F. 2016. Whole genome sequencing of Mycobacterium tuberculosis for detection of recent transmission and tracing outbreaks: A systematic review. Tuberculosis 98:77–85.

Roetzer A, Diel R, Kohl TA, Rückert C, Nübel U, Blom J, Wirth T, Jaenicke S, Schuback S, Rüsch-Gerdes S, Supply P, Kalinowski J, Niemann S. 2013. Whole genome sequencing versus traditional genotyping for investigation of a Mycobacterium tuberculosis outbreak: a longitudinal molecular epidemiological study. PLoS Med 10:e1001387.

Role and value of whole genome sequencing in studying tuberculosis transmission. 2019.. Clin Microbiol Infect 25:1377–1382.

Stucki D, Brites D, Jeljeli L, Coscolla M, Liu Q, Trauner A, Fenner L, Rutaihwa L, Borrell S, Luo T, Gao Q, Kato-Maeda M, Ballif M, Egger M, Macedo R, Mardassi H, Moreno M, Tudo Vilanova G, Fyfe J, Globan M, Thomas J, Jamieson F, Guthrie JL, Asante-Poku A, Yeboah-Manu D, Wampande E, Ssengooba W, Joloba M, Henry Boom W, Basu I, Bower J, Saraiva M, Vaconcellos SEG, Suffys P, Koch A, Wilkinson R, Gail-Bekker L, Malla B, Ley SD, Beck H-P, de Jong BC, Toit K, Sanchez-Padilla E, Bonnet M, Gil-Brusola A, Frank M, Penlap Beng VN, Eisenach K, Alani I, Wangui Ndung’u P, Revathi G, Gehre F, Akter S, Ntoumi F, Stewart-Isherwood L, Ntinginya NE, Rachow A, Hoelscher M, Cirillo DM, Skenders G, Hoffner S, Bakonyte D, Stakenas P, Diel R, Crudu V, Moldovan O, Al-Hajoj S, Otero L, Barletta F, Jane Carter E, Diero L, Supply P, Comas I, Niemann S, Gagneux S. 2016. Mycobacterium tuberculosis lineage 4 comprises globally distributed and geographically restricted sublineages. Nat Genet 48:1535–1543.

Surie D, Fane O, Finlay A, Ogopotse M, Tobias JL, Click ES, Modongo C, Zetola NM, Moonan PK, Oeltmann JE. n.d. Molecular, Spatial, and Field Epidemiology Suggesting TB Transmission in Community, Not Hospital, Gaborone, Botswana - Volume 23, Number 3—March 2017 - Emerging Infectious Diseases journal - CDC. doi:10.3201/eid2303.161183

The transmission of Mycobacterium tuberculosis in high burden settings. 2016. Lancet Infect Dis 16:227–238.

Walker TM, Lalor MK, Broda A, Ortega LS, Morgan M, Parker L, Churchill S, Bennett K, Golubchik T, Giess AP, Del Ojo Elias C, Jeffery KJ, Bowler ICJW, Laurenson IF, Barrett A, Drobniewski F, McCarthy ND, Anderson LF, Abubakar I, Thomas HL, Monk P, Smith EG, Walker AS, Crook DW, Peto TEA, Conlon CP. 2014. Assessment of Mycobacterium tuberculosis transmission in Oxfordshire, UK, 2007-12, with whole pathogen genome sequences: an observational study. Lancet Respir Med 2:285–292.

Whole-genome sequencing to delineate Mycobacterium tuberculosis outbreaks: a retrospective observational study. 2013.. Lancet Infect Dis 13:137–146.

Wood DE, Salzberg SL. 2014. Kraken: ultrafast metagenomic sequence classification using exact alignments. Genome Biol 15:1–12.

Xu Y, Cancino-Muñoz I, Torres-Puente M, Villamayor LM, Borrás R, Borrás-Máñez M, Bosque M, Camarena JJ, Colomer-Roig E, Colomina J, Escribano I, Esparcia-Rodríguez O, Gil-Brusola A, Gimeno C, Gimeno-Gascón A, Gomila-Sard B, González-Granda D, Gonzalo-Jiménez N, Guna-Serrano MR, López-Hontangas JL, Martín-González C, Moreno-Muñoz R, Navarro D, Navarro M, Orta N, Pérez E, Prat J, Rodríguez JC, Ruiz-García MM, Vanaclocha H, Colijn C, Comas I. 2019. High-resolution mapping of tuberculosis transmission: Whole genome sequencing and phylogenetic modelling of a cohort from Valencia Region, Spain. PLoS Med 16:e1002961.

Yang C, Lu L, Warren JL, Wu J, Jiang Q, Zuo T, Gan M, Liu M, Liu Q, DeRiemer K, Hong J, Shen X, Colijn C, Guo X, Gao Q, Cohen T. 2018. Internal migration and transmission dynamics of tuberculosis in Shanghai, China: an epidemiological, spatial, genomic analysis. Lancet Infect Dis 18:788–795.

